# Analysis of the functioning of temporary dialysis catheters in patients with COVID-19

**DOI:** 10.1101/2024.10.14.24314426

**Authors:** Bruno Jeronimo Ponte, Viviane Galli Dib, Felipe Soares Oliveira Portela, Arthur Souza Magnani, Marcela Juliano Silva, Lucas Lembrança Pinheiro, Nelson Wolosker

## Abstract

2.

**Introduction:** Patients with acute renal failure requiring hemodialysis should use temporary hemodialysis catheters (THC) due to the urgency and potential reversibility of the condition. So far, three studies in North America indicate a higher risk of catheter-related issues in COVID-19 patients needing hemodialysis. This study examines the functionality and complications of temporary hemodialysis access in COVID-19 patients at a Brazilian hospital.

**Methodology:** A prospective analysis was conducted at a COVID-19 referral center between May and July 2020. During this time, the Vascular Surgery team implanted temporary hemodialysis catheters in 107 patients. The patients were followed, and demographic and clinical characteristics that could be correlated with catheter malfunction were analyzed.

**Results:** Of the 107 patients studied, 22 (20,6%) experienced complications related to the catheter. Eighteen (16,8%) had malfunctions, and 4 (3,7%) had infections. Five patients evolved with unfavorable clinical conditions and did not undergo catheter replacement. Thirteen patients with malfunctions had catheter tip thrombosis as the cause of the malfunction. Among the variables analyzed, only the need for orotracheal intubation *(p 0,009)*, deep vein thrombosis *(p 0,01)*, and a history of a previous catheter *(p 0,002)* were found to be correlated with a higher risk of malfunctioning.

**Conclusion:** The rate of temporary high-flow catheter malfunction in patients with COVID-19 is similar to that in patients without this disease. Previous catheter use, the necessity for OTI, and venous thrombosis were the main predictors of catheter malfunction.

## 3. Introduction

The coronavirus disease 2019 (COVID–19) was first documented in Wuhan, China, in early December 2019. It was declared a pandemic by the World Health Organization (WHO) in March 2020. (1) The first case in Brazil was reported in February 2020 in Sao Paulo. One year later, there were about 9.4 million cases in Brazil, and the country was the 3rd in the world in the number of cases. (2,3)

COVID-19 can cause a range of symptoms, from mild to severe. While most cases are not severe, (4) there is a high risk of thrombosis affecting both the arteries and veins, especially in severe cases admitted to the ICU. (5) The infection also increases the risk of acute kidney injury (AKI) and the need for hemodialysis. AKI further raises thromboembolic risk. (6) For patients requiring hemodialysis due to acute renal failure, establishing vascular access with temporary hemodialysis catheters (THC) is crucial due to the condition’s potential reversibility and the urgency of treatment. (7)

Complications can occur following the implantation of THC, including infection, dislodgement, and malfunction due to thrombosis. (8,9) To date, only three North American studies have shown an increased risk of catheter-related malfunction in patients with COVID-19 under hemodialysis. (10–12)

Retrospective analyses by Kanitra et al., Shanmugasundaram et al., and Ouyang et al. evaluated hemodialysis catheter malfunction in COVID-19 patients. Kanitra et al. reported 31.3% malfunction in 48 patients, Shanmugasundaram et al. found 23.4% in 64 patients, and Ouyang et al. identified 22.6% malfunction due to thrombosis in 109 patients. (10–12)

Although these studies have shown a high incidence of malfunction of THC in patients with COVID-19 and AKI, no other studies were found involving different populations around the world, including Latin America.

This study aimed to analyze the functioning and complications of temporary hemodialysis accesses in a Brazilian hospital for a COVID-19-infected population.

## 4. Casuistry and Methods

In this observational and retrospective study conducted at a secondary referral hospital during the COVID-19 pandemic from April to July 2020, 107 THC were inserted in 107 patients with acute renal failure requiring hemodialysis due to COVID-19 infection.

Our institution’s Ethics Committee approved the study protocol and review board and conducted following the Declaration of Helsinki. It was registered under the number 39458720.8.0000.0071 at the Brazilian Platform Ethics.

Table 1 presents the demographic and technical characteristics of the patients. The patients’ ages ranged from 43 to 84 (58,2 ± 14,75). Most were male (64.5%). The body mass index (BMI) ranged 19,6 to 69,3kg/m^2^ (30,9± 7,9kg/m^2^). Twenty-six patients (24.3% %) required orotracheal intubation (OTI) at some point, and the majority (67.2% %) had some degree of hemodynamic instability requiring vasoactive drugs.

**Table 1.** Demographic and technical characteristics of patients.

| Variant | Average / (n) | % |
| --- | --- | --- |
| Age | 58,2 ± 14,75 (43-84) |  |
| Sex |  |  |
| Men | 69/107 | 64,5% |
| Women | 38/107 | 35,5% |
| BMI | 30,9 ± 7,9kg/m<sup>2</sup> (19,6-69,3) |  |
| OTI | 26/107 | 24,3% |
| Hemodinamic Instability | 72/107 | 67,2% |
| Puncture Site |  |  |
| Jugular | 93/107 | 86,9% |
| Femoral | 14/107 | 13,1% |
| D-Dimer | 4421 ± 2691ng/mL (462-8000ng/mL) |  |

*BMI: body mass index; OTI: orotracheal intubation*

A vascular surgery team member carried out the implantation procedures at the bedside. Local anesthesia was used, and all punctures were guided by ultrasound. The THC used were Biomedical ® Polyurethane Double Lumen catheters, with sizes of 11Fr x 15cm, 11Fr x 20cm, and 12Fr x 25cm. The majority of patients (86,9%) had catheters placed in the internal jugular vein (IJV), while 14 (13,1%) had them inserted into the common femoral vein (CFV).

After the catheter was implanted, the flow and reflux of both routes were tested to ensure they were functioning properly. Before use, an X-ray confirmed that the catheter was positioned at the cavo-atrial junction, right atrium, or inferior vena cava.

After the procedure, sterile dressings were applied to the puncture site. The catheters were filled with a diluted unfractionated heparin and saline solution (NaCl 0.9%) at 50 IU/mL, following the manufacturer’s recommended volume for each pathway.

The catheters showed adequate flow and reflux in all procedures and were cleared for use in hemodialysis. Once implanted, the system was immediately ready for use. No complications related to the device implantation were documented during or after the procedure.

During patient follow-up, the nursing team changed the dressing daily using an aseptic technique and monitored for signs of inflammation at the catheter site.

Patients were followed up for 7 days, and the follow-up was shortened for those who died.

The primary outcome analyzed was the incidence of THC-related complications that required device removal, including malfunction and infection. Malfunction was defined as the inability to perform hemodialysis due to the impossibility of aspiration and injection through the catheter routes. Infection was a presumed diagnosis since the patients had significant clinical severity, and was considered when patients presented with new onset of fever and chills associated with hemodynamic instability during hemodialysis sessions.

The study’s secondary outcome was identifying risk factors associated with catheter malfunction. The variables accessed included BMI, need for OTI, hemodynamic instability requiring vasoactive drugs, anticoagulation, catheter positioning, deep vein thrombosis, and D-Dimer levels.

To assess deep vein thrombosis (DVT), the vascular surgery team performed daily Doppler ultrasound evaluations of the venous system associated with the puncture site.

Statistical analysis comparing the groups was conducted using Fisher’s exact test for categorical variables and Student’s t-test for continuous variables. A p-value of less than 0.05 was considered statistically significant for all tests. The SPSS 20.0 Windows program (IBM Corp, Arnonk, NY) was used to conduct the analyses.

## 5. Results

Of the 107 catheters implanted, 22 (20.6%) had some symptomatic complications related to the device. Eighteen (16.82%) malfunctioned, and four (18.2%) had an infection. Table 2 describes the complications.

**Table 2.** Catheter related symptomatic complications.

| Variant | *n* | % |
| --- | --- | --- |
| Complications | 22 | 20,6% |
| Malfunction | 18 | 16,8% |
| Infection | 4 | 3,7% |
| Catheter Replacement | 17 | 15,8% |
| No Clinical Conditions for Replacement | 5 | 4,6% |

Out of 18 malfunctions, 13 (12.14%) were attributed to obstruction by thrombosis, where a clot was found at the tip of the catheter during device replacement and was removed and replaced. The remaining 5 patients experienced the same issue and required catheter replacement, but they were not in a clinical condition to undergo the procedure and passed away during follow-up.

The four patients with catheter infections presented symptoms of bacteremia during hemodialysis sessions, which were associated with worsening hemodynamics, fever, and chills. Therefore, the catheters were removed and replaced.

Thirty-eight patients (35.51%) had deep vein thrombosis associated with the catheter. Of these, 34 had sufficient clinical conditions and were fully anticoagulated. The other patients did not receive anticoagulation once their risk for bleeding was high.

We found no statistically significant correlation between age *(p 0,87)* or BMI *(p 0,45)* and catheter malfunction. Similarly, there was no correlation between catheter malfunction and access site *(p 0*.*292)* or hemodynamic instability *(p 0*.*10)*. However, we found significant correlations between catheter malfunction and a history of previous catheterization *(p 0,002)* and with clinical severity related to OTI *(p 0,009)*.

Regarding venous thrombosis, 38 (35,51%) out of 107 patients had DVT. There was a slightly significant correlation DVT and malfunction *(p 0*.*01)*. Although there was a correlation between DVT and catheter failure, patients who were on full anticoagulation did not correlate with lower catheter failure rates *(p 0*.*628)*

The mean D-Dimer value found in the sample was 4421ng/mL ± 2691 (*462-8000ng/mL*) and showed no correlation with catheter malfunction or venous thrombosis *(p 0*.*09)*. Only 62 out of 107 patients had D-Dimer collected during follow-up.

## 6. Discussion

Patients with COVID-19 are at greater risk of experiencing clinical complications involving both venous and arterial thrombosis. (13) They are also more likely to experience acute renal dysfunction requiring hemodialysis, especially in more severe patients. (14)

Severely ill patients who develop AKI and need hemodialysis have the implantation of THC as their first option. Device-related complications are more frequent in this population compared to non-critical patients. (11,15–18) In our sample, we found that patients with signs of clinical severity, including the need for OTI, were at greater risk of hemodialysis catheter malfunction *(p 0*.*009)*.

The reasons for this are based on the greater inflammatory cascade in these patients, leading to a greater risk of thromboembolic complications. (17,19) Added to the thrombogenic factor of COVID-19 and AKI itself with the need for hemodialysis, we observed that these patients had a greater relation with catheter malfunction.

Data on patients without COVID-19, under hemodialysis, after AKI with THC have a similar risk for catheter thrombosis and malfunction compared to our analysis. In a retrospective study of 77 implanted THCs, Jones et al. described a 23.37% rate of malfunction and an 11.68% rate of catheter-related thrombosis. (20) Additionally, Ouyang et al. analyzed the population without COVID-19 who were under hemodialysis and found that 8,2% dialysis access malfunction, including catheters and arteriovenous fistulas. (11)

In contrast to patients on hemodialysis for chronic kidney disease, without COVID-19 and using long-term catheters, they have about a 7.1% rate of catheter malfunction. (21)

The malfunction rate found in our population (16.8%) was similar to that found in North American studies evaluating THC in patients with AKI and COVID-19. Between 22.6% and 31.3% of the patients analyzed had malfunctioned catheters. (10–12)

Elderly and obese patients are at greater risk of COVID-19-related complications, including thrombosis and acute renal failure. (22–24) In our analysis, we found no relationship between age and catheter malfunction. Other studies evaluating the risk of catheter thrombosis in hemodialysis COVID-19 patients have also not identified advanced age as a risk factor for catheter malfunction. (10–12)

In a retrospective analysis, Ouyang et al. analyzed THC function in COVID-19 patients and found that there was a higher ratio of THC malfunction in obese patients (*mean BMI 33*.*1; p<0*.*001)*. (11)

Although most of the patients assessed in our analysis were obese, no factors associated with higher risks of catheter malfunction or infection were identified in this population.

The preferred access route for implanting most of the THC in our service is via puncture of the internal jugular vein, which was successfully utilized in 93 of our patients (86,9%). This vein provides a safe route directly to the right atrium. Its punction requires minimal tissue manipulation and, when guided by ultrasound, allows a safe and simple procedure, avoiding possible complications. (25)

For catheters placed in the internal jugular veins, positioning the tip at the cavo-atrial or even intra-atrial junction is recommended, as this location reduces the formation of a fibrin cap. (18,26)

In femoral catheters, the recommended positioning for the catheter tip is in the inferior vena cava, as it provides adequate flow and reflux through the pathways, which is usually achieved in catheters longer than 24cm. (25,27)

When we evaluated the relation between device positioning and risk of malfunction, we found no significant correlation between the variables *(p 0,37)*.

In our analysis, the presence of previous central venous access was correlated with THC malfunction. As is well known, endothelial injury caused by catheter implantation and morphological changes in the vessel wall due to local trauma from the insertion of an intravascular device may contribute to subsequent catheter failure. (18) Hence, preserving puncture sites before catheter implantation is important, especially in patients on chronic hemodialysis programming.

Of the 22 patients with complications related to the device, 13 had malfunction related to thrombi at the tip of the catheter, with flow obstruction. Analyzing the 38 patients with deep vein thrombosis related to the puncture site, we found a correlation between thrombosis and catheter malfunctioning *(p 0,01)*.

These data corroborate those found in the literature, where patients with severe conditions and a higher risk of thrombosis have higher rates of catheter thrombosis, leading to malfunction. (28,29)

The prophylactic measures established and standardized by the service included heparinizing the catheter routes with an unfractionated heparin solution. Studies have identified a lower chance of catheter thrombosis when heparinization of hemodialysis catheter routes is performed, compared to full anticoagulation and prophylactic subcutaneous dosing in the COVID-19 and hemodialysis population. (12,30)

Finally, even with high levels of D-dimer, this marker was not related to the prevalence of catheter malfunction and/or venous thrombosis, confirming that, despite its role in predicting general prognosis, it is not directly useful concerning catheter use. This finding was corroborated by Ouyang et al., in which no correlation was observed between D-Dimer and THC function. (11)

Our study had several limitations. Other comorbidities and patients’ epidemiological data were not assessed, which could lead to selection bias since other diseases can increase the risk of thrombosis. In addition, much information could not be collected, and D-Dimer was only evaluated in 62 of the 107 patients.

Also, by evaluating the first seven days after catheter implantation, it was impossible to analyze devices that could have malfunctioned after this period, reducing the

Despite this, we present a study evaluating catheter function in COVID-19 patients with AKI and hemodialysis in the largest sample to date and the first carried out in a population outside the United States.

## 7. Conclusion

The rate of temporary high-flow catheter malfunction in patients with COVID-19 is similar to that in patients without this disease. Previous catheter use, the necessity for OTI, and venous thrombosis were the main predictors of catheter malfunction.

## Data Availability

All data produced in the present work are contained in the manuscript.

